# Identifying implementation units that would benefit from alternative treatment strategies to accelerate the elimination of onchocerciasis transmission in Africa

**DOI:** 10.64898/2026.07.02.26357114

**Authors:** Aditya Ramani, Matthew A. Dixon, Martin Walker, Raiha Browning, Evandro Konzen, Simon E. F. Spencer, Claudio Fronterre, Maria-Gloria Basáñez

## Abstract

**Background:** The World Health Organization proposes that elimination of onchocerciasis transmission (EOT) be verified in 12 endemic countries by 2030. In sub-Saharan Africa (SSA), where most cases occur, Niger is the only country that has been verified to date. Despite decades of ivermectin mass drug administration (MDA), infection persists in West and Central Africa. Alternative treatment strategies (ATS) are necessary to accelerate progress towards EOT by 2030 and beyond.

**Methods:** We used the EPIONCHO-IBM transmission model to project the number of years, from 2026, to reduce microfilarial (mf) prevalence below 1% across 1,634 implementation units (IUs) in 19 SSA countries. We fitted the model to geostatistically-derived mf prevalence in 1975, 2000 and 2018, and projected mf prevalence through to 2025. We classified IUs according to their baseline endemicity, intervention history programmatic performance, and current (2025) MDA frequency (annual or biannual). For those IUs that would not reach < 1% mf prevalence by 2030 if current strategies were continued, we simulated ATS (increasing treatment frequency, improving coverage, and adopting moxidectin MDA) from 2026 to 2040.

**Results:** Of the 1,486 IUs currently under annual ivermectin MDA, 45% would require ATS. In those low-moderate endemicity IUs, biannual ivermectin would have a comparable impact to that of switching to annual moxidectin; in those with high endemicity, adopting biannual moxidectin would be more impactful. Of the 148 IUs currently receiving biannual ivermectin, 24% would benefit from ATS, switching to biannual moxidectin being the best option.

**Conclusion:** This work brings into sharper focus which IU profiles are most likely to require ATS across SSA. In highly-endemic IUs with long intervention histories, biannual moxidectin MDA may be required under our modelling assumptions, with substantial uncertainty surrounding the permanent sterilising effect, of the two drugs under comparison, upon adult female worms. National programmes aiming to reach EOT will have options that need to be balanced against financial considerations. Implementation studies will also be important for translating these modelling projections into national policy decisions, particularly where different intervention strategies generate similar epidemiological benefits. Economic and epidemiological evaluations of repeated moxidectin MDA are needed to inform these decisions.

## 1 Introduction

Onchocerciasis (‘river blindness’), a vector-borne neglected tropical disease (NTD), is caused by infection with the filarial nematode *Onchocerca volvulus*, affecting communities located in river basins in endemic countries, where the blackfly *Simulium* spp. vectors breed. More than 99% of all onchocerciasis cases are concentrated in sub-Saharan Africa (SSA), where, in 2024, a total population of 251 million required mass drug administration (MDA) with ivermectin, and a reported 172 million were treated (1). Onchocerciasis is associated with substantial morbidity due to skin and eye disease, contributing 1.26 (95% uncertainty interval, UI = 0.75–1.90) million disability-adjusted life-years (DALYs) according to the 2021 Global Burden of Disease (GBD) study (2). In addition to cutaneous and ocular clinical sequelae (including blindness), onchocerciasis is associated with epilepsy (3), and responsible for excess mortality, attributed to blindness (4), increasing *O. volvulus* microfilarial (mf) load (5), and epilepsy (6).

Two major regional initiatives have shaped onchocerciasis control efforts across SSA, namely, the Onchocerciasis Control Programme in West Africa (OCP, 1974-2002), and the African Programme for Onchocerciasis Control (APOC, 1995-2015) (7, 8). National NTD programmes have subsequently continued control efforts, with technical assistance from the Expanded Special Project for Elimination of Neglected Tropical Diseases (ESPEN). During the OCP, large-scale and prolonged vector control through weekly aerial larviciding of *S. damnosum* sensu lato (s.l.) breeding sites was implemented (9). During APOC, aerial larviciding was also conducted on the island of Bioko (Equatorial Guinea), leading to the elimination of the Bioko form of *S. yahense* (10), and ground larviciding against *S. neavei* was deployed in Uganda (11). In addition to or instead of vector control, ivermectin MDA was implemented in the OCP, initially through mobile teams of drug distributors from the late 1980s (7). APOC introduced community-directed treatment with ivermectin (CDTI) in the late 1990s (12), which was also adopted by the OCP. Annual ivermectin MDA has been the most widespread treatment strategy, but biannual (6-monthly) or more frequent treatment has also been implemented in Benin, Burkina Faso, Ethiopia, Ghana, Guinea, Guinea-Bissau, Nigeria, Senegal, Sierra Leone, South Sudan, Sudan, Tanzania, Togo and Uganda (13).

In 2021, the World Health Organization (WHO) Roadmap on NTDs proposed that 12 endemic countries be verified for elimination of onchocerciasis transmission (EOT) by 2030 (14). However, despite decades of intervention, only 8.5% of foci have reported EOT across SSA (15) and Niger is, to date, the only country verified by the WHO as having eliminated onchocerciasis (16). Equatorial Guinea and Senegal have stopped MDA and currently are under post-treatment surveillance (1). The prevalence of onchocerciasis continues to be concentrated across West and Central Africa. In 2018, focal areas of Angola, Cameroon, Democratic Republic of the Congo, Ethiopia, Ghana, Guinea, Mali, Nigeria, South Sudan and Uganda still had mean infection prevalence estimates greater than 25% (17). It is therefore clear that alternative treatment strategies (ATS) are necessary to accelerate the elimination of onchocerciasis by 2030 and beyond. ATS include, among others, increasing the frequency of ivermectin MDA and deploying new anthelmintics (18).

One such ATS would be the use of moxidectin, a macrocyclic lactone in the same drug class as ivermectin but with considerably longer half-life in humans (20-43 days compared to approximately 1 day for ivermectin) (19), resulting in more prolonged mf clearance from the skin following a single dose, as shown in Phase II and III clinical trials (20, 21, 22). The enhanced mf suppression effected by moxidectin would greatly reduce the potential for transmission of microfilariae from humans to vectors between treatment rounds, suggesting that annual moxidectin MDA could be as effective as biannual ivermectin MDA (23).

In 2018, the US Food and Drug Administration (FDA) approved moxidectin for the treatment of *O. volvulus* infection in persons aged 12 years and older (19). Following the results of a study aimed at finding a paediatric dose of moxidectin for the treatment of 4-11-year-olds (24), in 2025 the US FDA expanded its indication for the treatment of persons as young as 4 years (25). The Ghana Food and Drugs Authority had approved its marketing authorization in 2024 (26), and in 2025, the WHO included moxidectin among its list of essential medicines (27). Ghana, having been the first country where a community trial of ivermectin took place in 1987-1992 in the highly hyperendemic focus of Asubende in the Pru district (28), is now the first country in SSA where moxidectin MDA is being implemented biannually in an area of the Twifo Atti Morkwa district, with ongoing transmission despite many years of prior ivermectin treatment (29).

A previous, scenario-based modelling study had suggested that biannual moxidectin MDA could reduce by half the number of years necessary to achieve EOT in mesoendemic areas and might be the only strategy that can achieve it in hyperendemic areas (30). Sub-national planning for onchocerciasis control and elimination efforts is currently conducted at the implementation unit (IU) level across SSA. To support countries more widely in decisions regarding the adoption of moxidectin MDA as an ATS, this modelling analysis expands our previous work (30) to identify which IUs would benefit the most from continuing current treatment strategies, increasing the frequency and/or coverage of ivermectin MDA if not already done so, or rolling out community-based treatment with moxidectin and which frequency and coverage should be implemented to reach < 1% mf prevalence by 2040. We used the value of less than 1% mf prevalence as an indication of being close to EOT following Mutono et al. (15).

## 2 Methods

### 2.1 EPIONCHO-IBM

EPIONCHO-IBM is a stochastic, individual-based model (31), which extends a previous deterministic framework (32). Within individual humans, the model tracks the number of *O. volvulus* adult (male and female) worms and that of microfilariae per milligram of skin (or per skin snip). Female worms comprise non-fertile and fertile females to account for the fact that juvenile females are initially non-fertile and that mature females need 3-4 re-insemination cycles per year to produce microfilariae, cycling between these two states (33). The dynamics of microfilariae within humans as well as of (L1, L2, L3) larvae in the vector are modelled deterministically. Exposure to blackfly bites depends on human age and sex (32). Additional inter-individual exposure heterogeneity, represented by parameter *k_E_* (31), gives rise to overdispersed distributions of parasite burden among hosts that are the hallmark of helminth infections. Parasite population abundance is regulated by density-dependent processes assumed to operate upon parasite establishment within humans and vectors, vector survival, and the probability that a female worm is mated. For each value of *k_E_* (0.2, 0.3, 0.4)—inversely related to the magnitude of exposure heterogeneity—there is an associated set of density dependence parameters determining the within-humans parasite establishment probability. This probability is a decreasing function of transmission intensity (the number of infective, L3 larvae/person/year) (31). It is further assumed that there is a balanced sex ratio of male and female worms and that the parasites are polygamous (33). A detailed description of the model can be found in Hamley et al. (31). Supplementary File 1, Supplementary Figure 1 provides a schematic representation of EPIONCHO-IBM.

### 2.2 Modelling interventions

MDA was modelled assuming that a proportion of eligible individuals receive treatment with a given drug. For ivermectin, eligible individuals are those aged ≥ 5 years; for moxidectin, those aged ≥ 4 years (25). A microfilaricidal effect and a temporary embryostatic effect were modelled using expressions and parameters previously described for ivermectin (34) and moxidectin (23). Additionally, a permanent sterilizing (anti-macrofilarial) effect per treatment dose was assumed to operate from the second round of treatment onwards (23, 30, 31). By way of sensitivity analysis, the anti-macrofilarial effect was taken to result in a 35% or 70% irreversible reduction in the number of fertile females. The former value followed a previous estimation for ivermectin (35) used also for moxidectin in previous modelling studies (30). The latter value (henceforth referred to as MOX MAX) was used to reflect the longer half-life of moxidectin compared to ivermectin (19) as well as uncertainty surrounding this effect. Table 1 presents the approximate estimated microfilaridermia reduction after 1 and 52 weeks following single-dose treatment with either drug and the assumptions made regarding the permanent sterilizing effect. Supplementary File 1, Supplementary Methods 1 and Supplementary Table 1 provide the expressions and parameter values used to model the effects of ivermectin and moxidectin on *O. volvulus*.

**TABLE 1.** Efficacy assumptions for ivermectin and moxidectin.

|  | Approximate net reduction (%) of microfilaridermia (microfilaricidal and embryostatic effects) following treatment |  | Permanent sterilization effect (from second dose) |
| --- | --- | --- | --- |
|  | After 1 week | After 52 weeks |  |
| Ivermectin (IVM) | 94% | 80% | 35% |
| Moxidectin (MOX) | 100% | 98% | 35% |
| Moxidectin max (MOX MAX) | 100% | 98% | 70% |

Adherence to treatment was modelled following Dyson et al. (36) and as previously done in EPIONCHO-IBM (37). To this end, a parameter (*ρ*) is selected between the values of 0 (random adherence: eligible individuals are treated at random and have the same chance of receiving treatment at any given MDA round) and 1 (fully systematic adherence: the same group of eligible individuals is treated at every round and the remainder are never treated). For the purposes of our study, *ρ* = 0.3 was used, reflecting data on the number of rounds attended during ivermectin MDA for Burundi (38), and Cameroon and Nigeria (39).

### 2.3 Implementation units (IUs) and intervention histories

Both baseline (pre-intervention) onchocerciasis endemicity levels and intervention histories used for all the IUs modelled in this study were derived from a precursor version of the HISTONCHO dataset for onchocerciasis control and elimination in SSA (13). With the earliest interventions starting with vector control in 1974-75 in the OCP, HISTONCHO collates data from ESPEN (40), regional and country reports, and other literature to reconstruct intervention histories across onchocerciasis-endemic IUs in SSA. In this analysis, IUs were categorized according to their baseline endemicity (reflecting infection prevalence prior to the implementation of intervention(s), Table 2), and historical programmatic performance as per ESPEN records (Table 3). IUs identified as under post-intervention surveillance according to ESPEN were not considered for this analysis (3 IUs in Ethiopia, 32 in Nigeria, and 8 in Uganda). Countries already verified for EOT (Niger), under post-treatment surveillance (Equatorial Guinea and Senegal), deemed not to require MDA (Kenya, Rwanda) (41), or not in ESPEN (Sudan) were not included. Additionally, IUs where no MDA data were available in ESPEN, but other reference sources indicated that some intervention(s) may have been implemented in years before ESPEN records begun (2013) were also excluded due to uncertainty surrounding intervention history. A total of 1,671 IUs were eligible for inclusion in the analysis. HISTONCHO uses ESPEN data up to 2022, and infers MDA status for 2023-2025, including whether IUs remain on annual or biannual treatment frequency (13).

**TABLE 2.**
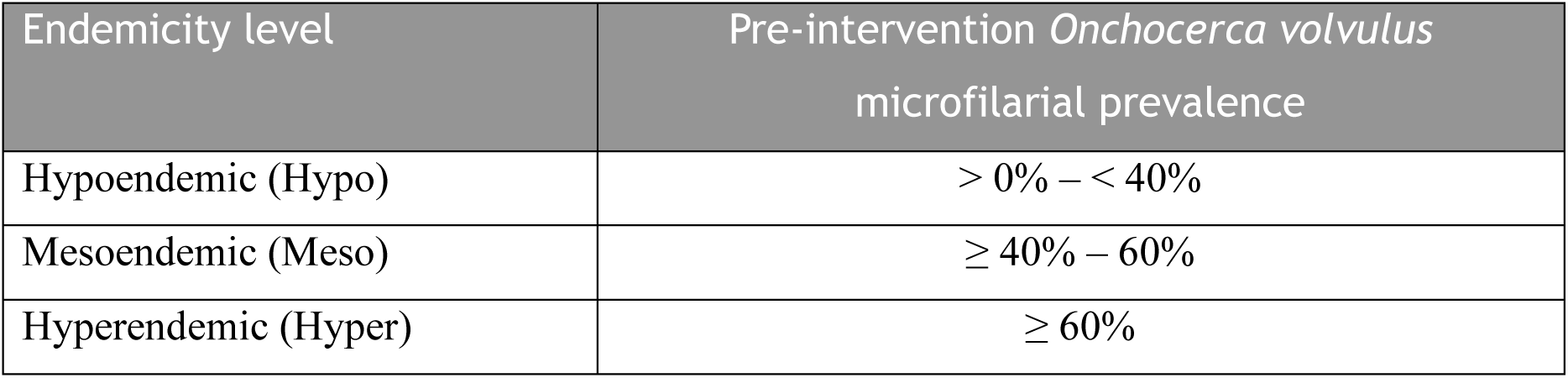
Classification of baseline endemicity levels.

| Endemicity level | Pre-intervention *Onchocerca volvulus* microfilarial prevalence |
| --- | --- |
| Hypoendemic (Hypo) | > 0% – < 40% |
| Mesoendemic (Meso) | ≥ 40% – 60% |
| Hyperendemic (Hyper) | ≥ 60% |

**TABLE 3.** Classification of intervention history programmatic performance.

| Performance | Criteria |  |
| --- | --- | --- |
|  | Number of consecutive years of MDA<sup>a</sup> in the ESPEN<sup>b</sup> database | Coverage of total population<sup>c</sup> (all ages) |
| High | ≥ 8 years | All years with ≥ 65% coverage |
| Medium | ≥ 8 years | Some years with < 65% coverage |
| Low | < 8 years | All years with ≥ 65% coverage |
| Lowest | < 8 years | Some years with < 65% coverage |
| Treatment naïve | No previous MDA according to ESPEN/HISTONCHO up to 2022 |  |

<sup>a</sup> MDA: Mass drug administration, in this case of ivermectin.
<sup>b</sup> ESPEN: Expanded Special Project for Elimination of Neglected Tropical Diseases; the ESPEN database contains MDA data from 2014-2022 (13).
<sup>c</sup> A coverage of 65% of total population is the minimum coverage for onchocerciasis control (48) and is approximately equivalent to a coverage of 80% of eligible population.

### 2.4 Fitting EPIONCHO-IBM to multiple-time point data and simulating intervention histories to 2025

To fit the model to the mf prevalence temporal trends in each IU, a modified version of the adaptive multiple importance sampling (AMIS) algorithm (42) was used. In brief, a Bayesian inference approach based on importance sampling was used to fit the annual biting rate (ABR = the number of blackfly vector bites/person/year) and the individual exposure heterogeneity (*k_E_*) parameters that determine mf prevalence in EPIONCHO-IBM (31). These parameters were fitted to three data points, namely, 1975 (i.e. baseline/pre-intervention), 2000 and 2018. The data for each time point were based on pixel-level mf prevalence maps generated using model-based geostatistics (MBG) from: (i) O’Hanlon et al. (43) for the OCP area at baseline, (ii) Zouré et al. (44) for APOC countries at baseline, and (iii) Schmidt et al. (17) for 2000 and 2018 across SSA. Since the MBG-generated prevalence for APOC countries at baseline was based on nodule (onchocercomata) prevalence in adult male samples aged ≥ 20 years (44), its conversion into mf prevalence in those aged ≥ 5 years was implemented (45, 46). The pixel-level mf prevalence data were aggregated into a prevalence distribution for each IU at each time point. Simulations for each parameter set were run to the final time point (2018) using previously determined priors on ABR and *k_E_* centred around reasonable values determined by other literature (31). The likelihood for a given parameter set for each IU was defined as the probability that the EPIONCHO-IBM-modelled mf prevalence was drawn from the prevalence distribution for that IU, aggregated as a cumulative likelihood across all three time points.

An effective sample size (47) was calculated with weights derived from the likelihoods derived as described above and compared to a target of 500. Once the target effective sample size was reached, 200 simulations (200 ABR and *k_E_* combinations) were subsampled for each IU, using the weights obtained from the AMIS algorithm. These weights represented how well each parameter set aligned with mf prevalence. These initial simulations were projected from baseline endemicity through to the final time point of 2018. From 2019, simulations based on each of the 200 subsampled parameter sets were continued according to the intervention histories recorded in HISTONCHO (13) for each IU through to 2025.

### 2.5 Identifying IUs under current MDA strategies that would require ATS

The IU mf prevalence was simulated (using 200 parameter sets as described above) from 2026 through to 2030 on the basis of continuing its current MDA strategy (annual or biannual ivermectin MDA), assuming a 65% coverage of total population. Those IUs that were projected not to reach < 1% mf prevalence by 2030 were considered for simulating different ATS. Figure 1 presents the breakdown of IUs in each baseline endemicity and intervention performance category in 2025, their current MDA strategy, and the number of IUs projected not to reach < 1% mf prevalence by 2030 if their current intervention strategy were continued from 2026. Figure 1 also includes a map indicating the geographical distribution of IUs currently under annual ivermectin, biannual ivermectin, and projected to reach < 1% mf prevalence by 2030 if continuing the current strategy. For a map presenting the IU distribution of historical interventions, see (13). Of the 1,671 IUs, 31% were hypoendemic at baseline, 65% were mesoendemic and 4% were hyperendemic, with 37 (2%) being treatment-naïve (97% of which were hypoendemic at baseline). Twenty-seven treatment-naïve IUs (73%) were located in Gabon, all onchocerciasis hypoendemic but co-endemic with hyperendemic loiasis (13). In these IUs, it is unlikely that routine MDA will be implemented. This is due to the risk of severe adverse events (SAE) following microfilaricidal treatment of highly *Loa loa* microfilaraemic individuals (18). Therefore, Gabon was excluded from our analysis. The remaining treatment-naïve IUs were in Ethiopia (7), Nigeria (2) and Togo (1), which were also excluded. Of the 1,486 IUs currently under annual ivermectin MDA, 815 (55%) would reach < 1% mf prevalence by 2030. Of the 148 IUs currently under biannual ivermectin MDA, 76% would attain this target.

**FIGURE 1.**
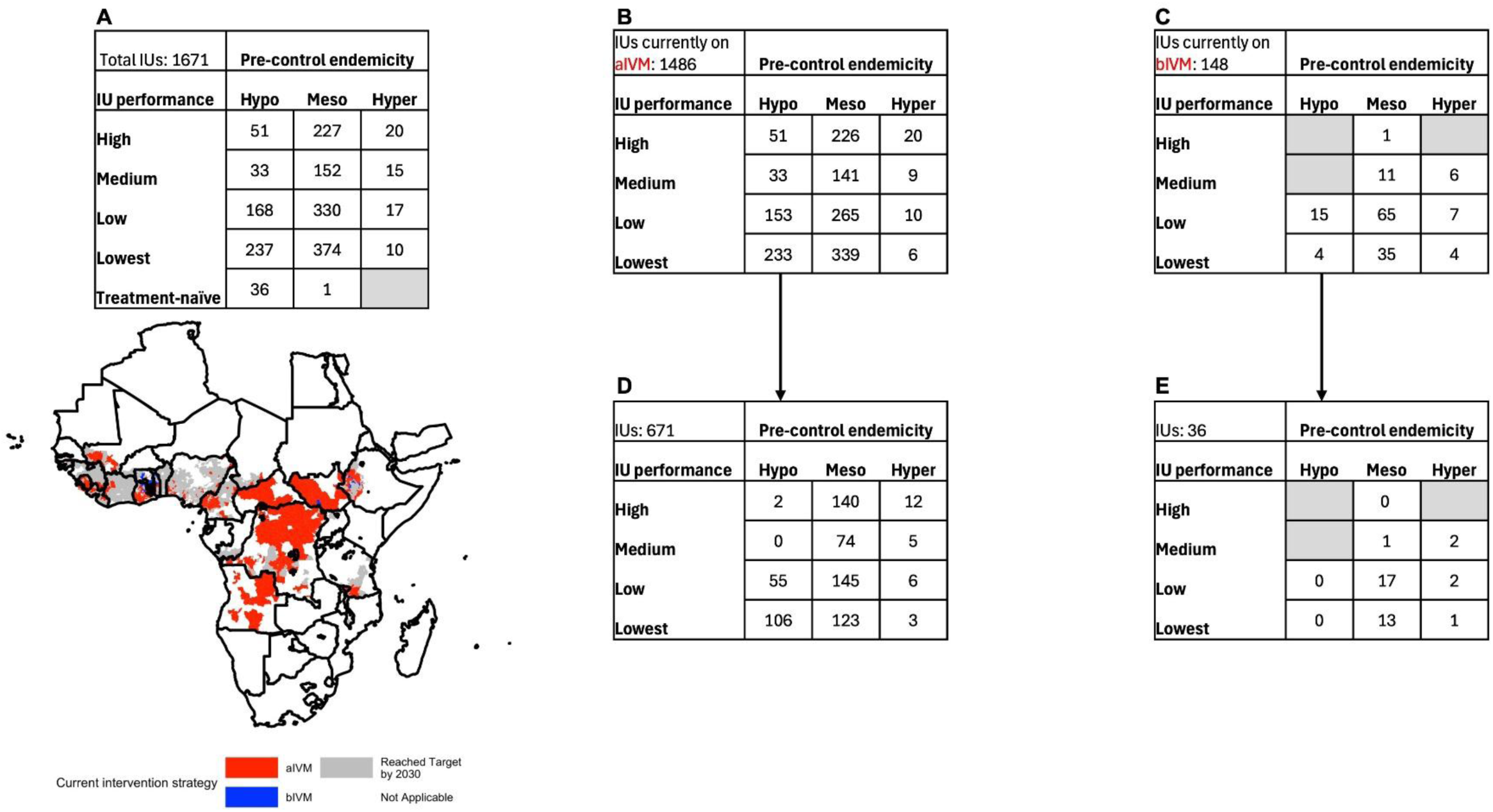
Onchocerciasis-endemic implementation units (IUs) classified according to their baseline (pre-intervention) endemicity, intervention history performance, and current intervention strategy. (**A**) Total IUs eligible for this work and map of their location in sub-Saharan Africa according to current intervention strategy: annual ivermectin (aIVM, red); biannual ivermectin (bIVM, blue); projected to reach < 1% *Onchocerca volvulus* microfilarial (mf) prevalence by 2030 if continuing current (aIVM or bIVM) strategy (grey); not applicable (white); (**B**) IUs undergoing annual ivermectin mass drug administration (MDA) in 2025 (aIVM); (**C**) IUs undergoing biannual ivermectin MDA in 2025 (bIVM); (**D**) IUs currently on aIVM that would not reach < 1% mf prevalence by 2030 if the current strategy were continued; (**E**) IUs currently on bIVM that would not reach < 1% mf prevalence by 2030 if the current strategy were continued. For a map indicating the IU distribution of historical interventions, see (13). Grey cells in the tables of **(A)**, **(C)** and **(E)** indicate absence of IUs in the particular combination. IU pre-control endemicity: Hypoendemic (Hypo), Mesoendemic (Meso), Hyperendemic (Hyper) (Table 2); IU performance: High, Medium, Low, Lowest (Table 3).

### 2.6 Simulating ATS

In those IUs not attaining < 1% mf prevalence by 2030, ATS were simulated from 2026 to 2040 (200 model runs for each combination). Each combination comprises a choice of: drug (ivermectin or moxidectin), coverage (standard coverage: 65% of total population or enhanced coverage: 80% of total population), and frequency (annual or biannual). The 65% coverage (approximately 80% coverage of eligible population) has been considered as the minimum necessary to control onchocerciasis, whereas 80% coverage (nearly 100% coverage of eligible population) would be necessary to achieve EOT (48). Table 4 presents a matrix of ATS for the 12 combinations. We calculated the number of years necessary to reach < 1% mf prevalence by the end of 2040, across all IUs in each one of the countries analysed. The number of years was defined as the mean of all 200 simulations for each IU and each of the 12 ATS combinations (Supplementary File 1, Supplementary Methods 2).

**TABLE 4.** Alternative treatment strategy (ATS) combinations.

| Frequency<sup>a</sup> | Coverage of total population<sup>b</sup> | Drug<sup>c</sup> |  |  |
| --- | --- | --- | --- | --- |
|  |  | Ivermectin (IVM) | Moxidectin (MOX) | Moxidectin max (MOX MAX) |
| Annual | 65% (Standard) | A \| S IVM | A \| S MOX | A \| S MOX MAX |
|  | 80% (Enhanced) | A \| E IVM | A \| E MOX | A \| E MOX MAX |
| Biannual | 65% (Standard) | B \| S IVM | B \| S MOX | B \| S MOX MAX |
|  | 80% (Enhanced) | B \| E IVM | B \| E MOX | B \| E MOX MAX |

<sup>a</sup> Frequency is denoted by A for annual and B for biannual (6-monthly) mass drug administration (MDA).
<sup>b</sup> Coverage of total population (all ages) is denoted by S for standard coverage (65%; corresponding approximately to 80% of eligible population) and E for enhanced coverage (80%; corresponding to nearly 100% of eligible population). A total population coverage of 80% is recommended for elimination of onchocerciasis transmission (48).
<sup>c</sup> For MOX MAX, the permanent sterilising effect is assumed to be 70%, twice as high as the 35% estimated for ivermectin (35), per dose (Table 1, and Supplementary File 1, Supplementary Table 1).

We followed the five principles of the NTD Modelling Consortium regarding Policy-Relevant Items for Reporting Models in Epidemiology of NTDs (PRIME-NTD) (49) (Supplementary File 1, Supplementary Table 2).

## 3 Results

Compared across the same frequencies and coverages, modelling projections suggest that moxidectin would outperform ivermectin through its greater, faster and more sustained declines in mf prevalence. Figure 2 illustrates this with one high prevalence IU (in Togo) and one low prevalence IU (in Tanzania), also highlighting that when the prevalence is already low in 2026, the difference between the treatment strategies is less pronounced.

**FIGURE 2.**
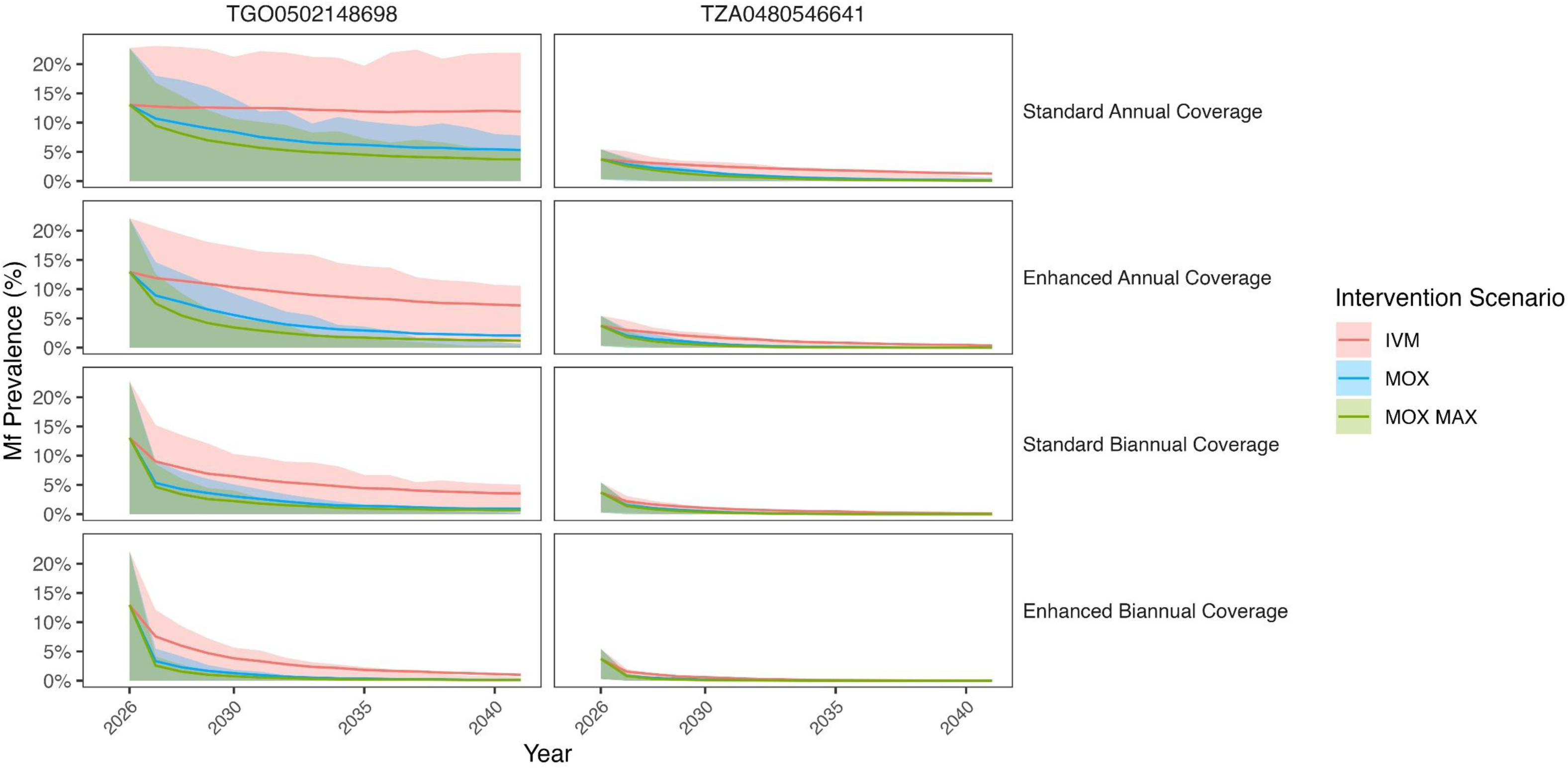
Impact of alternative treatment strategies (ATS) on *Onchocerca volvulus* microfilarial (mf) prevalence (%) trajectories (from 2026 to 2040) according to EPIONCHO-IBM projections. **(A)** High prevalence implementation unit (IU) in Togo (TGO0502148698); **(B)** low prevalence IU in Tanzania (TZA0480546641). Lines show the median mf prevalence and shaded areas the 95% uncertainty intervals (2.5^th^-97.5^th^ quantiles) of 200 stochastic simulations for: ivermectin (IVM, red); moxidectin (MOX, blue); moxidectin max (MOX MAX, green). The rows denote coverage and frequency of mass drug administration (MDA): standard coverage (65% of total population) and annual frequency (top row: Standard Annual Coverage); enhanced coverage (80% of total population) and annual frequency (second row: Enhanced Annual Coverage); standard coverage and biannual frequency (third row: Standard Biannual Coverage); enhanced coverage and biannual frequency (bottom row: Enhanced Biannual Coverage).

### 3.1 Distribution across IUs of the number of years to attain < 1% mf prevalence

The distributions of the (mean) number of years necessary to reach < 1% mf prevalence by 2040, across all IUs in each one of the 19 countries analysed are shown in Figure 3. In Burundi, Burkina Faso, Malawi and Uganda, all IUs were projected to have reached < 1% mf prevalence before 2026. Therefore, these countries are not included. In Ethiopia, Ghana, Nigeria, South Sudan, Tanzania and Togo biannual ivermectin MDA had already been implemented in several IUs before 2026 (13). However, to facilitate comparison between treatment strategies, annual ivermectin MDA was also simulated for all IUs in these countries.

**FIGURE 3.**
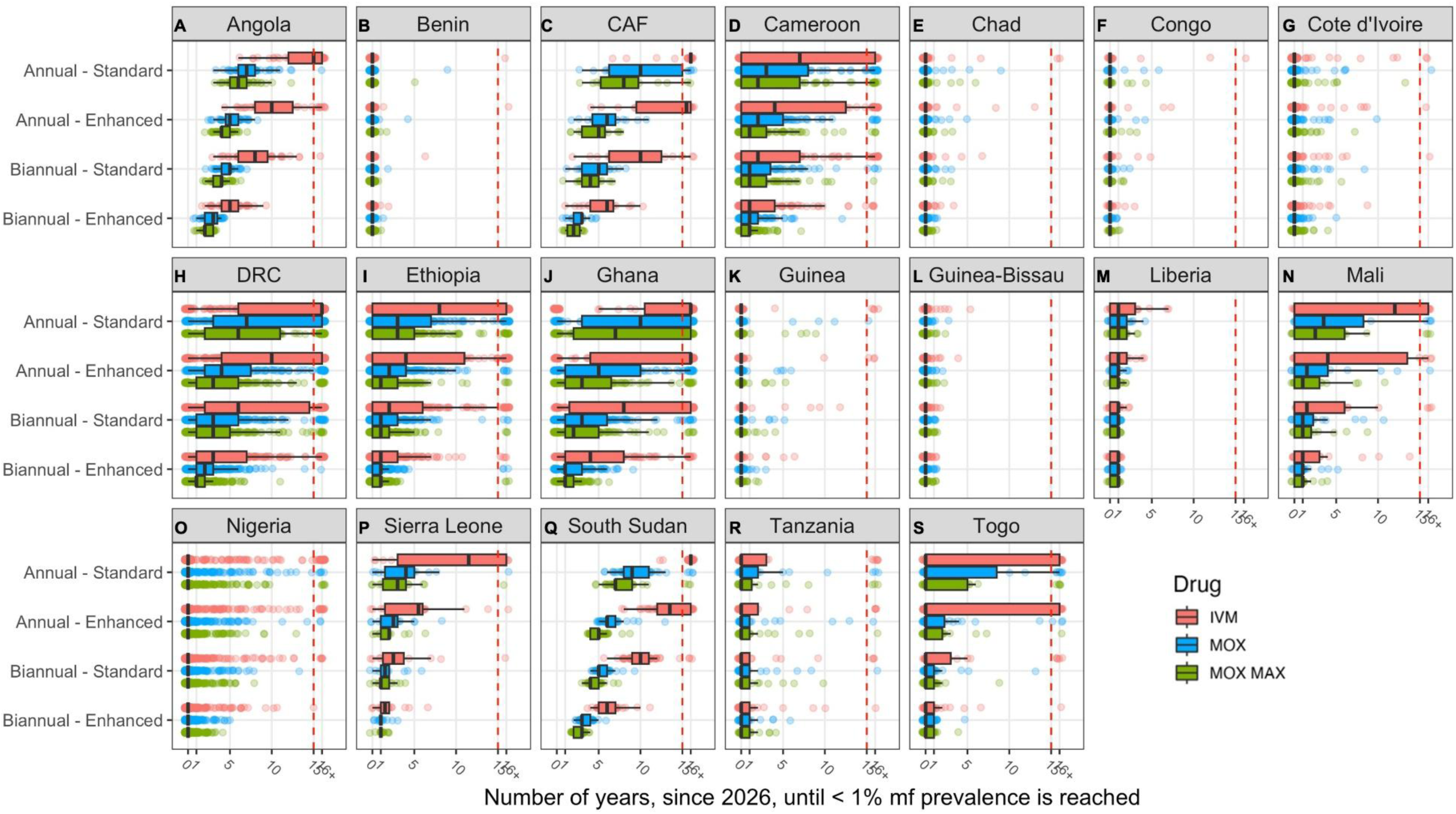
Distribution of the (mean) number of years necessary to reach < 1% *Onchocerca volvulus* microfilarial (mf) prevalence across implementation units (IUs) for 19 countries in sub-Saharan Africa according to EPIONCHO-IBM. Boxplots show the median (black vertical line within the box), interquartile range (IQR, edges of the box), and 1.5 times the IQR (whiskers of the box) number of years to reach < 1% mf prevalence across all IUs for a given country. The colours of the circles (which represent each IU within the country) and the boxplots denote: ivermectin (IVM, red); moxidectin (MOX, blue); moxidectin max (MOX MAX, green). **(A)** Angola; **(B)** Benin; **(C)** Central African Republic (CAF); **(D)** Cameroon; **(E)** Chad; **(F)** Congo; **(G)** Côte d’Ivoire; **(H)** Democratic Republic of the Congo (DRC); **(I)** Ethiopia; **(J)** Ghana; **(K)** Guinea; **(L)** Guinea-Bissau; **(M)** Liberia); **(N)** Mali; **(O)** Nigeria; **(P)** Sierra Leone; **(Q)** South Sudan; **(R)** Tanzania; **(S)** Togo. The simulated interventions are: annual mass drug administration (MDA) frequency with standard coverage (65% of total population): Annual - Standard; annual MDA frequency with enhanced coverage (80% of total population): Annual - Enhanced; biannual MDA with standard coverage: Biannual - Standard, and biannual MDA with enhanced coverage: Biannual - Enhanced. The vertical red dashed line indicates the last year of treatment simulated (2040, 15 years of MDA from 2026). Those IUs not reaching < 1% mf prevalence by the end of 2040 are represented by circles to the right of the vertical dashed red line (16+ years).

In Cameroon, where the current strategy is annual ivermectin, switching to (standard coverage) annual moxidectin would reduce the upper quartile number of years to reach < 1% mf prevalence from more than 16 to 8 (> 50% reduction), and adopting biannual moxidectin would result in a 78% reduction (Figure 3D), with further reductions projected under enhanced coverage. In Ghana, where the current strategy is mostly biannual ivermectin, implementing biannual moxidectin would reduce the upper quartile number of years to reach < 1% mf prevalence from more than 16 to 6 (> 63% reduction) under standard coverage or to 3 years (> 81% reduction) under enhanced coverage (Figure 3J). In South Sudan, annual ivermectin MDA with standard coverage would result in only two IUs reaching < 1% mf prevalence by 2040. Annual moxidectin with standard coverage would lead to over 75% of IUs reaching the target (Figure 3Q).

### 3.2 IUs currently under annual ivermectin MDA

EPIONCHO-IBM projections suggest that, of the 1,486 IUs under annual ivermectin MDA at the time of our modelling study (Figure 4A), 815 (55%) would reach < 1% mf prevalence by 2030, but 671 (45%) would not, with the majority of these IUs (508, 76%) classified as mesoendemic or hyperendemic at baseline, and 277 (41%) categorized as having a poor (‘low’ or ‘lowest’) intervention history performance (Figure 4B). The remaining panels of Figure 4 refer to standard coverage only. Continuing with annual frequency of ivermectin MDA through to 2040 would only result in 186 IUs (28%) reaching < 1% mf prevalence (Figure 4C). Switching to biannual ivermectin MDA would increase this number to 537 (80%) (Figure 4D), with a similar outcome when adopting annual moxidectin MDA (499 IUs, 74%, or 552, 82% under hypothetical moxidectin max) (Figures 4E, 4G). Switching to biannual moxidectin MDA would increase the number of IUs reaching < 1% mf prevalence to 644 (96%) or to 658 (98%) if moxidectin had twice as high a permanent sterilizing effect as that assumed for ivermectin (70% vs. 35%) (Figures 4F, 4H).

**FIGURE 4.**
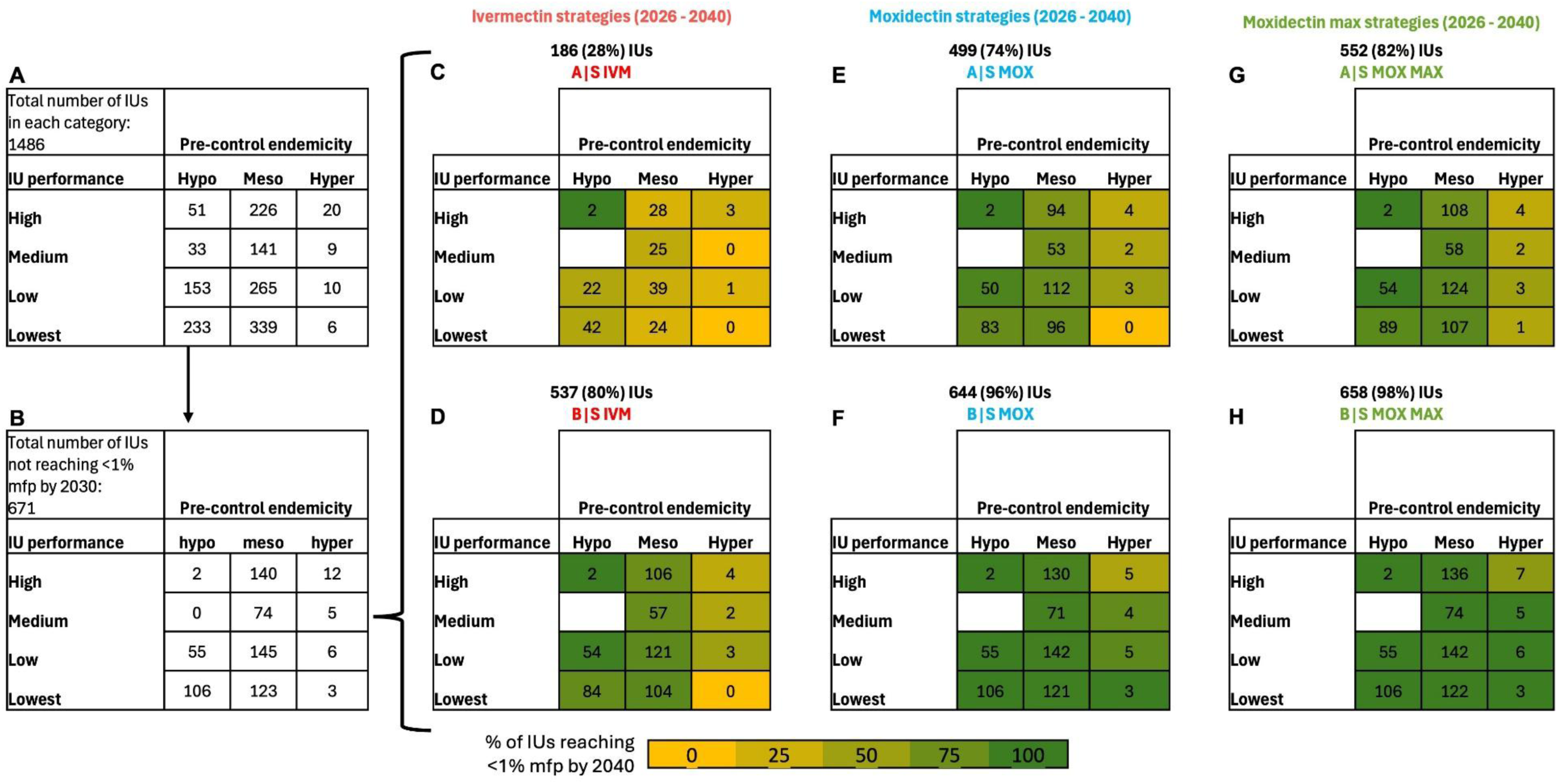
Implementation units (IUs) currently under annual ivermectin mass drug administration (MDA), those which would not reach < 1% *Onchocerca volvulus* microfilarial prevalence (mfp) by 2030 under the same strategy, and what number of IUs would reach it by the end of 2040 under standard coverage (S, 65% of total population) treatment strategies (Table 4) simulated from 2026. IUs are classified by pre-control endemicity (Hypoendemic, Mesoendemic, Hyperendemic) and programmatic performance (High, Medium, Low, Lowest). **(A)** Total number of IUs currently implementing annual ivermectin MDA; **(B)** IUs that would not reach < 1% mfp by 2030 if continuing annual ivermectin; **(C)** IUs that would reach < 1% mfp by 2040 if continuing annual ivermectin MDA (A | S IVM); **(D)** IUs that would reach < 1% mfp by 2040 if switching to biannual ivermectin (B | S IVM); **(E)** IUs that would reach < 1% by 2040 if switching to annual moxidectin (A | S MOX); **(F)** IUs that would reach < 1% mfp by 2040 if switching to biannual moxidectin (B | S MOX); **(G)** IUs that would reach < 1% mfp by 2040 if switching to annual moxidectin under the assumption that its permanent sterilizing effect is twice that of ivermectin (A | S MOX MAX); **(H)** IUs that would reach < 1% mfp by 2040 if switching to biannual moxidectin under the assumption that its permanent sterilizing effect is twice that of ivermectin (B | S MOX MAX). Cells indicate the number of IUs classified according to baseline endemicity (Table 2) and historical programmatic performance (Table 3). The colour of the cells indicates the proportion of IUs projected to reach < 1% mfp by 2040, from 0% (dark yellow) to 100% (dark green). White cells in **(C)** to **(H)** are blank as the corresponding number of IUs (33) would have reached < 1% mfp by 2030 **(A, B)**

### 3.3 IUs currently under biannual ivermectin MDA

Of the 148 IUs currently under biannual ivermectin MDA (Figure 5A), 112 (76%) are projected to reach < 1% mf prevalence by 2030, but 36 (24%) would not do so, all being meso- or hyperendemic at baseline and 92% with poor (low/lowest) historical programmatic performance (Figure 5B). The remaining panels of Figure 5 refer to standard coverage only. Of the 36 IUs, 10 (28%) would reach < 1% mf prevalence by continuing biannual ivermectin MDA through to 2040 (Figure 5C). Returning to annual frequency but switching to moxidectin would not be beneficial (only 4-9, 11-25%, IUs would attain < 1% mf prevalence) by 2040 (Figures 5D, 5F) while adopting biannual moxidectin is projected to result in 29-30 (81-83%) of IUs reaching the target (Figures 5E, 5G).

**FIGURE 5.**
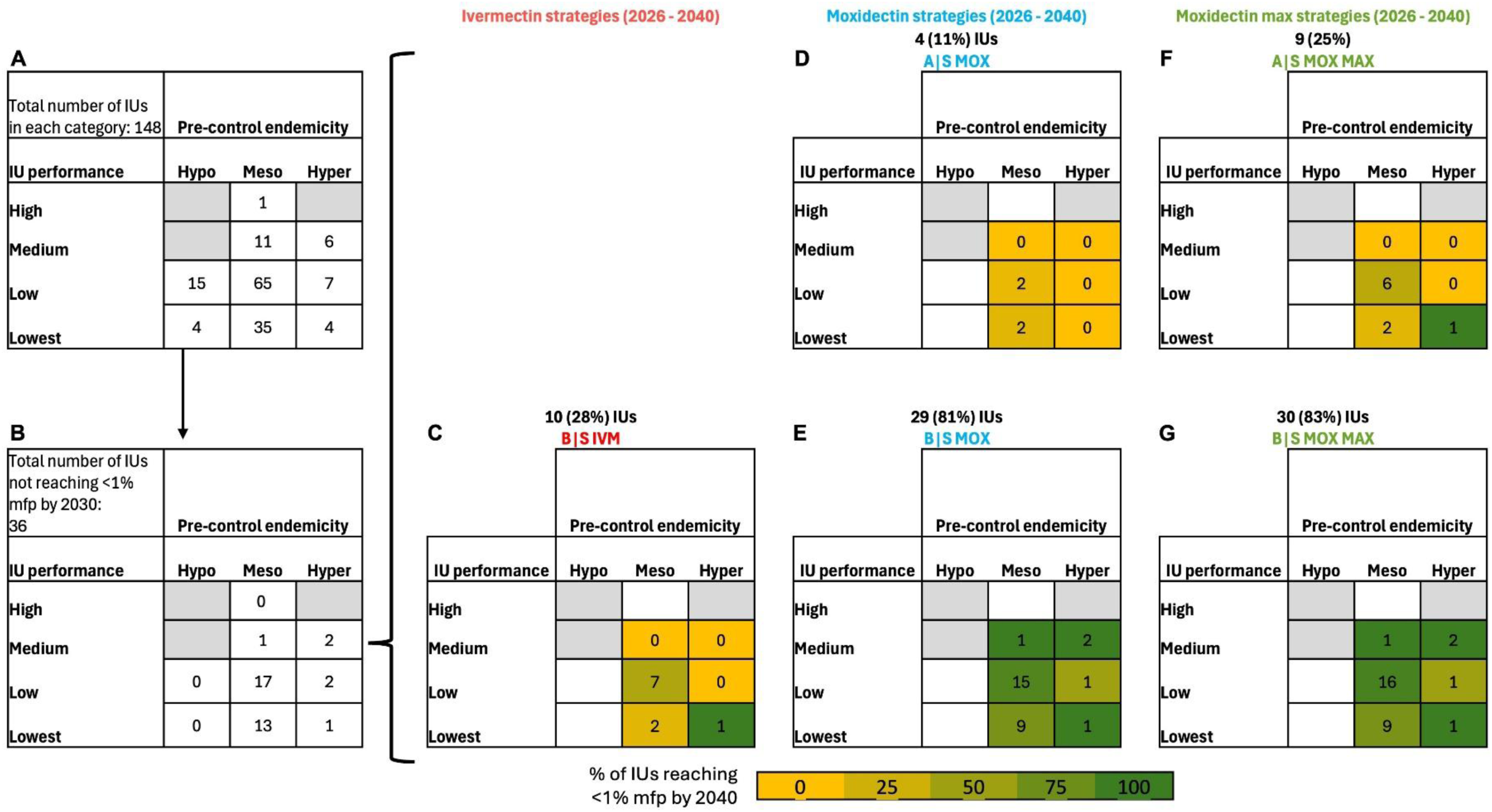
Implementation units (IUs) currently under biannual ivermectin mass drug administration (MDA), those which would not reach < 1% *Onchocerca volvulus* microfilarial prevalence (mfp) by 2030 under the same strategy, and what number of IUs would reach it by the end of 2040 under standard coverage (S, 65% of total population) treatment strategies (Table 4) simulated from 2026. IUs are classified by pre-control endemicity (Hypoendemic, Mesoendemic, Hyperendemic) and programmatic performance (High, Medium, Low, Lowest). **(A)** Total number of IUs currently implementing biannual ivermectin MDA; **(B)** IUs that would not reach < 1% mfp by 2030 if continuing biannual ivermectin; **(C)** IUs that would reach < 1% mfp by 2040 if continuing biannual ivermectin MDA (B | S IVM); **(D)** IUs that would reach < 1% mfp by 2040 if switching to annual moxidectin (A | S MOX); **(E)** IUs that would reach < 1% mfp by 2040 if switching to biannual moxidectin (B | S MOX); **(F)** IUs that would reach < 1% mfp by 2040 if switching to annual moxidectin under the assumption that its permanent sterilizing effect is twice that of ivermectin (A | S MOX MAX); **(G)** IUs that would reach < 1% mfp by 2040 if switching to biannual moxidectin under the assumption that its permanent sterilizing effect is twice that of ivermectin (B | S MOX MAX). Cells indicate the number of IUs classified according to baseline endemicity (Table 2) and historical programmatic performance (Table 3). The colour of the cells indicates the proportion of IUs projected to reach < 1% mfp by 2040, from 0% (dark yellow) to 100% (dark green). Grey cells indicate absence of IUs with a given combination of pre-control endemicity and programmatic performance. White cells in **(C)** to **(G)** are blank as the corresponding number of IUs (1) would have reached < 1% mfp by 2030 **(A, B)**.

## 4 Discussion

The modelling results presented in this study are the first to consider the impact of ATS across onchocerciasis-endemic IUs in SSA using data-informed baseline endemicities, intervention trajectories since their initiation and historical programmatic performance (13). Therefore, we extend our previous scenario-based modelling work (30), providing an updated epidemiological situation up until 2025 as well as the projected number of years to reduce mf prevalence below 1% by 2040 in 19 SSA countries. We used this threshold to indicate being close to EOT (15), but do not provide probabilities of reaching EOT (30). Our aim was to identify which combination of IU characteristics would indicate the need to deploy ATS to accelerate EOT in the run up to 2030 and beyond. We did not model local vector control or test-and-treat/not-treat strategies, which are also considered among ATS (18). Our analysis specifically focused on where it would be advantageous (from a purely epidemiological perspective) to increase treatment frequency and/or coverage of ivermectin MDA, compared to the introduction of moxidectin under two contrasting assumptions regarding its potential irreversible sterilizing effect upon female worms. Thus, we do not consider economic implications, as it is yet uncertain what financial mechanisms will operate in different countries if moxidectin, a not donated drug, were to be adopted in certain IUs where ivermectin MDA has proven insufficient.

Based on the results of our previous scenario-based modelling study (30), Turner et al. (50) concluded that moxidectin-based strategies could not only accelerate progress towards EOT but also reduce programmatic delivery costs compared with ivermectin-based strategies. These authors emphasized, however, that obtaining data on the costs of moxidectin to national programmes would be essential to quantify whether delivery cost reductions will translate into overall programme cost reduction (50).

In agreement with Turner et al. (23) and Kura et al. (30), our results (Figure 3) suggest that the epidemiological impact of annual moxidectin MDA is similar to that of biannual ivermectin. However, in areas where biannual ivermectin is already ongoing, continuing with biannual ivermectin could be more impactful than switching to annual moxidectin, but less so than switching to biannual moxidectin.

Our projections indicate that, for IUs with hypo- or mesoendemic baseline endemicity currently under annual ivermectin, switching to annual moxidectin would result in 75% or more IUs reaching < 1% mf prevalence by 2040 across all intervention performance characteristics, compared with 50% or less achieving this if annual ivermectin MDA were continued (with the exception of those IUs with high historical programmatic performance). Switching to biannual ivermectin would lead to a somewhat greater but comparable change in the proportion of IUs achieving mf prevalence below 1% by 2040. This finding would provide countries with flexibility to choose between increasing treatment frequency to biannual ivermectin and switching to annual moxidectin to achieve similar results. Decisions will be based on the costs to their programme of implementing biannual ivermectin (51, 52) vs. the costs of annual moxidectin (50).

In those IUs that were hyperendemic at baseline, especially those where programmatic performance had been poor (low/lowest), our modelling results suggest that switching to biannual moxidectin would be preferable to continuing with annual ivermectin, contingent on coverage being improved (to reach at least 80% of eligibles) and consistently maintained over MDA rounds.

There were fewer IUs (148 compared to 1,486) currently under biannual ivermectin MDA, reflecting that the predominant strategy across SSA has been annual treatment. Only 36/148 (24%) were projected not to reach < 1% mf prevalence by 2030. This (modelling) result agrees with the finding that biannual ivermectin MDA was statistically significantly and positively associated with the odds of achieving EOT/being close to EOT in the systematic review and meta-analysis conducted by Mutono et al. (15). These authors found that only 7% of the records in their study reported implementation of biannual ivermectin, which is close to our 9% (148/1,634) across all analysed IUs. We found that a higher proportion of IUs was projected to reach the target by 2040 if biannual ivermectin were continued compared to reverting to an annual frequency but switching to moxidectin (28% vs. 11% or vs. 25% under hypothetical MOX MAX), particularly in IUs that had been at least mesoendemic at baseline and had poor (low/lowest) historical programmatic performance, which represented the great majority of IUs (Figure 5). The greatest benefit seemed to derive from maintaining biannual frequency but switching to moxidectin (81-83% of IUs attaining mf prevalence below 1% by 2040), under the proviso that coverage is improved (≥ 80% of eligible population) and sustained.

In countries with a prolonged intervention history which still have focal areas with high infection prevalence (e.g. Ghana, Mali and Togo, formerly under OCP, and Cameroon, Ethiopia and Nigeria, formerly under APOC), our projections suggest that adopting biannual moxidectin would be the most impactful strategy (Figure 3), particularly in IUs with high baseline endemicity. A modelling study using survey mf prevalence data (1970–2017) from 400 villages in Togo, reported that in highly hyperendemic areas at baseline, prevalence rebounded even under biannual ivermectin MDA, indicating the need for ATS in general and biannual moxidectin in particular (53).

The present study has several limitations that shall be addressed in future work. These limitations concern the consideration of < 1% mf prevalence as an endpoint; the assumption of 65% or 80% coverage of total population for the 2026 onwards simulations; the simulation of MDA in IUs where onchocerciasis and loiasis are co-endemic; the use of a restricted set of density dependence parameters that influence the rate at which mf prevalence increases during inter-treatment periods (resilience to MDA); the modelling of closed populations, and the uncertainty surrounding the permanent sterilizing effect assumed for ivermectin and moxidectin.

Reaching < 1% mf prevalence is an indication of substantial progress, but not necessarily a predictor of achieving EOT. Infection could resurge if MDA were stopped at that point, with the probability and rate of resurgence depending on baseline endemicity and prevailing transmission conditions (53). Modelling EOT at IU level would entail the modelling of pre-stop/stop-MDA surveys, as well as the dynamics of (serological and entomological) indicators during post-treatment and post-elimination surveillance, which is beyond the scope of this paper.

The coverage assumptions for the ATS period, comparing the impact of 65% or 80% of total population, are important determinants of the modelling results presented in Figure 2 and Figure 3, but obviously it is not possible to foresee the future coverage levels that will be attained and sustained in endemic IUs from 2026 onwards. For the results presented in Figure 4 and Figure 5 we opted for the more conservative assumption of 65% of total population (standard) coverage. The coverage values used for the 2026-2040 projections were informed by our previous modelling work using detailed reported coverage at subnational level (53).

Although we did not include Gabon in the analysis because of its widespread onchocerciasis-loiasis co-endemicity, we simulated MDA in IUs of other countries which face similar challenges (being endemic for *O. volvulus* infection as well as highly endemic for *L. loa* infection), such as Angola, Cameroon, Central African Republic, Congo, and Democratic Republic of the Congo (13). In these IUs, routine MDA is not advisable (18). Test-and-not-treat (TaNT) strategies—which test individuals for *L. loa* microfilaraemia and if this exceeds critical levels, do not provide treatment with microfilaricides—have been successfully piloted (54, 55), but we did not model TaNT in this study. Blok et al. (56), using the ONCHOSIM transmission model, undertook a scenario-based approach, simulating baseline *O. volvulus* mf prevalence values ranging from 2% to 40%, and varying levels of treatment adherence and exclusion from ivermectin treatment. These authors reported that implementing TaNT in treatment-naïve onchocerciasis hypoendemic areas co-endemic with loiasis, would require approximately 16-18 years to bring mf prevalence below 1.4% (56).

In areas co-endemic with loiasis, fear of SAE has resulted in low levels of treatment adherence and high proportions of systematic non-compliers (never-treated groups) (57) that hinder progress towards EOT (36). In meso- to hyperendemic IUs, where ivermectin MDA has already been implemented, the value of the *ρ* parameter would have to be increased to capture high proportions of systematic non-adherence (37). By contrast, in the TaNT pilot studies conducted in Cameroon, at least 60% of the total population presented for testing and 95% of those tested were safely treated in each round (54, 55). This would mean that after 5 years of annual TaNT, nearly all community residents would have been tested for loiasis at least once and, given that ivermectin treatment substantially reduces *L. loa* microfilaraemia for at least a year (58), it may be possible to follow 5 years of TaNT with implementation of MDA, at increased adherence. Modelling the impact of TaNT at IU level in onchocerciasis-loiasis co-endemic areas (13), is therefore an important (ongoing) area of work.

Although we fitted the ABR and *k_E_* parameters to the MBG-derived mf prevalence for three (1975, 2000, 2018) time points, we did not re-estimate the values of the density dependence parameters down-regulating within-humans parasite establishment as a function of transmission intensity. Instead, we used those corresponding to *k_E_* = 0.3 (31). In consequence, we may have underestimated the impact of treatment in hyperendemic IUs and overestimated it in hypoendemic IUs. In the former, exposure heterogeneity is likely to be less (greater *k_E_* values are associated with less severe density dependence, allowing EPIONCHO-IBM to generate higher mf prevalence but also yielding less resilience to interventions). In the latter, exposure heterogeneity may be stronger (smaller *k_E_* values are associated with more severe density dependence, stabilizing low mf prevalence but increasing resilience (31, 59).

We modelled closed populations and, therefore, our results must be interpreted in this context. However, immigration of infected individuals between IUs within countries, as well as between IUs (across borders) between neighbouring countries—which in both cases may differ in baseline endemicities, intervention histories and intervention performance characteristics—could be responsible for importation of infection and persistent transmission (60). This could result in longer timelines to reach < 1% mf prevalence than those projected here and potentially jeopardise EOT. Adopting a scenario-based approach, Stapley et al. illustrated this risk by using EPIONCHO-IBM to investigate the effect of infection importation from a highly endemic area into an infection-free area. The vulnerability of infection-free communities to introduction/re-introduction of infection depended on their population size and ABR, the number of immigrants arriving and their *O. volvulus* burden (60). The development of a spatially-structured version of EPIONCHO-IBM would be essential to understand the impact of human (and/or vector) movement on the IU based projections presented here and therefore constitutes a modelling priority.

Finally, substantial uncertainty surrounds the assumption of a permanent sterilizing effect of ivermectin (35), which we also used for moxidectin, as previously done (30). Other modelling studies do not support the operation of a cumulative effect of multiple ivermectin treatments on mf production by female worms (61), or estimate that multiple doses of ivermectin have a partial macrofilaricidal effect but only a modest permanent sterilizing effect (62). Turner et al. (23) varied the magnitude of this effect (1%, 7%, 30%) using a deterministic precursor of the stochastic version used here. With a 30% (and at times 7%) anti-macrofilarial action, biannual ivermectin MDA programmes were approximately equivalent in their epidemiological impact to annual moxidectin MDA programmes, but incurred higher total costs due to the increased treatment frequency (23). Kura et al. (30) conducted a sensitivity analysis halving the permanent sterilizing effect of ivermectin (from 35% to 17.5%) and assuming that the value of 35%, estimated by Plaisier et al. (35) for ivermectin, would apply to moxidectin. In the present study, we used the value of 35% for ivermectin and moxidectin, and doubled it for ‘MOX MAX’. Therefore, our projections must be interpreted with caution, as data are not yet available on the efficacy and effectiveness of multiple moxidectin doses (given annually or biannually) in the context of clinical trials (63) or large-scale MDA studies (29).

## 5 Conclusion

The adoption of the ATS investigated in this study (increasing treatment frequency of ivermectin MDA or switching to moxidectin MDA) holds promise to support national programmes of SSA onchocerciasis-endemic countries in their efforts to accelerate EOT by 2030 and beyond. However, this would not be a one-size-fits-all approach, but rather, a tailor-made decision that needs to be informed by IU-specific baseline endemicity, intervention history, programmatic performance, and the feasibility of improving and sustaining therapeutic coverage and treatment adherence. In IUs with low to moderate endemicity currently under annual ivermectin MDA, our modelling projections suggest that switching to annual moxidectin would lead to an epidemiological impact comparable to that of increasing treatment frequency to biannual ivermectin, but adopting biannual moxidectin would be more impactful in highly-endemic IUs. In IUs already under biannual ivermectin, switching to biannual moxidectin would represent the best option. These decisions will have to be made in the context of their financial repercussions to the national onchocerciasis control and elimination programmes. In addition to programme costs, the success of implementing MDA in general, and ATS in particular is highly dependent on contextual factors (64). The integration into MDA of a new drug like moxidectin requires securing national-level regulatory clearances (26), strengthening supply chains, expanding local health workforce capabilities, overcoming community hesitancy, and managing logistical hurdles in endemic areas (e.g. changing from height-pole dosing for ivermectin to tablet-dosing for moxidectin). Whilst awaiting evaluations regarding the cost, feasibility and acceptability of implementing moxidectin MDA, as well as the results of clinical and field trials assessing the efficacy and effectiveness of repeated moxidectin treatments, our work helps to bring into sharper focus where ATS are most needed across IUs in SSA.

## Data Availability

This work is a mathematical modelling study and, therefore, no human participants were recruited and no individual-level identifiable data were used. No institutional review board approval and informed consent were required. All data generated in the present work are contained in the manuscript. The HISTONCHO dataset that was used to inform the intervention histories and programmatic performance characteristics for the modelling is openly accessible at: https://zenodo.org/records/15390119. The code used to reconstruct the intervention histories is available at: https://github.com/mrc-ide/HISTONCHO. The code for the EPIONCHO-IBM model is available at: https://github.com/mrc-ide/EPIONCHO.IBM.

## Data availability statement

The HISTONCHO dataset comprises the data used to inform the intervention histories and programmatic performance characteristics used here and is openly accessible as RDS and CSV files at https://zenodo.org/records/15390119 (13).

## Code availability statement

The code used to reconstruct the intervention histories is available at: https://github.com/mrc-ide/HISTONCHO (13). The code for the EPIONCHO-IBM model is available at: https://github.com/mrc-ide/EPIONCHO.IBM.

## Ethics statement

For this study, no human participants were recruited and no individual-level identifiable data were used; therefore, no institutional review board approval and informed consent were required for the present (modelling) analysis.

## Authors’ contributions

Conceptualisation: A.R., M.A.D., M.W. and M.-G.B. Data curation: M.A.D., R.B. and C.F. Formal analysis: A.R., M.A.D., R.B., E.K. and C.F. Investigation and Resources: A.R., M.A.D., M.W., R.B., E.K., S.E.F.S., C.F. and M.-G.B. Methodology and Software: A.R., M.A.D., R.B., E.K., S.E.F.S. and C.F. Supervision and Funding acquisition: M.W., S.E.F.S. and M.-G.B. Visualisation: A.R., M.A.D., M.W. and M.G.B. Writing – original draft: A.R., M.A.D. and M.- G.B. Writing – review & editing: A.R., M.A.D., M.W., R.B., E.K., S.E.F.S., C.F. and M.-G.B.

## Acknowledgments

We thank Stefano Mavolti, Manuela Runge and Wilma Stolk for earlier discussions regarding use cases for moxidectin, and Igor Clark and Tom Eisner for their contribution to the IU modelling pipeline from 2026 to 2040. We also thank Andreia Vasconcelos (NTD Modelling Consortium secretariat) and Déirdre Hollingsworth for helpful feedback on previous versions of our results.

## Generative AI statement

The author(s) declare that no Generative AI was used in the creation of this manuscript.

## Publisher’s note

All claims expressed in this article are solely those of the authors and do not necessarily represent those of their affiliated organizations, or those of the publisher, the editors and the reviewers. Any product that may be evaluated in this article, or claim that may be made by its manufacturer, is not guaranteed or endorsed by the publisher.

## Supplementary material

**Supplementary File 1.** Supplementary Figure 1 (EPIONCHO-IBM model schematic); Supplementary Methods 1 (Modelling ivermectin and moxidectin treatment effects on *Onchocerca volvulus*); Supplementary Table 1 (Parameter definitions and values for anti-parasitic effects of ivermectin (IVM), moxidectin (MOX) and moxidectin max (MOX MAX) used in EPIONCHO-IBM); Supplementary Methods 2 (Calculation of the (mean) number of years to reach < 1% microfilarial prevalence); Supplementary Table 2 (Policy-Relevant Items for Reporting Models in Epidemiology of Neglected Tropical Diseases (PRIME-NTD) summary table); Supplementary References.

