## Supplementary File 1 for "Identifying implementation units that would benefit from alternative treatment strategies to accelerate the elimination of onchocerciasis transmission in Africa"

### Identifying implementation units that would benefit from alternative treatment strategies to accelerate the elimination of onchocerciasis transmission in Africa

Aditya Ramani[1,2§], Matthew A. Dixon[1,3§], Martin Walker[2], Raiha Browning[4], Evandro Konzen[4], Simon E. F. Spencer[4], Claudio Fronterre[5,6] and Maria-Gloria Basáñez[1*]

[1] MRC Centre for Global Infectious Disease Analysis, Department of Infectious Disease Epidemiology, School of Public Health, Imperial College London, 90 Wood Lane, London, W12 0BZ, UK

[2] Department of Pathobiology and Population Sciences, Royal Veterinary College, Hawkshead Lane, Hatfield, Hertfordshire, AL9 7TA, UK

[3] Unlimit Health (formerly known as SCI Foundation), Edinburgh House, 170 Kennington Lane, London, SE11 5DP, UK

[4] Department of Statistics, University of Warwick, Coventry, CV4 7AL, UK

[5] Centre for Health Informatics, Computing and Statistics (CHICAS), Lancaster University, Lancaster, LA1 4YW, UK

[6] Department of Applied Health Sciences, University of Birmingham, Edgbaston, Birmingham, B15 2TT, UK.

**Current addresses**

Raiha Browning: Walter and Eliza Hall Institute of Medical Research, 1G Royal Parade, Parkville, Melbourne, Victoria 3052 Australia

Evandro Konzen: HR Wallingford, Howbery Park, Benson Lane, Wallingford OX10 8BA, UK

[§]Contributed equally (joint first authors)

**Supplementary Figure 1.**

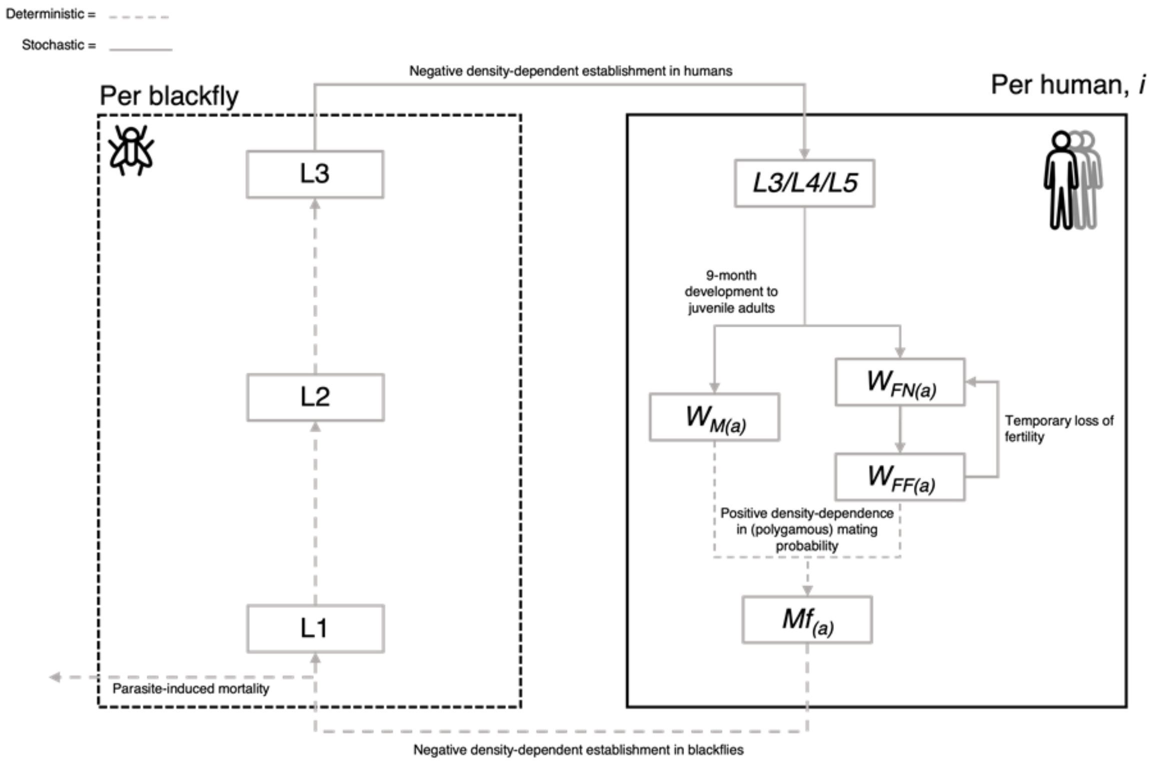


**EPIONCHO-IBM model schematic** (1)**.** EPIONCHO-IBM is an individual-based onchocerciasis transmission model (2), extending a deterministic precursor (3). The model considers *Onchocerca volvulus* infection dynamics in the blackfly vector (per blackfly box) and human hosts (per human box). Deterministic components are denoted by dashed lines; stochastic components by solid lines. Within an individual human host, *i*, the model tracks numbers of L3, L4, and L5 *O. volvulus* stages, adult worms (*W*) and microfilariae (*Mf*). Microfilariae are the L1 parasite stage. Numbers of adult males ($W_M$) and females ($W_F$) are tracked separately (assuming a 1:1 sex ratio). Females are initially non-fertile ($W_{FN}$) and cycle between fertile ($W_{FF}$) and non-fertile, reflecting their periodic reproduction biology (4). *Mf* are produced following the mating of adult male and fertile female worms, assuming complete polygamy (i.e., a single adult male can inseminate all fertile females). The survival of adult worms and microfilariae is modelled using a Weibull distribution, such that the average lifespan of adults is 9.28 years (with 95% dead by 12.0 years) and that of microfilariae is 0.833 years (with 95% dead by 1.09 years). Parasite fecundity decreases nonlinearly with age, the fecundity rate halving by 13 years (2). Individuals are exposed to blackfly bites depending on their age and sex (3), in addition to an individual exposure heterogeneity (gamma distributed) factor randomly assigned at birth (2). The latter leads to most individuals having low worm burdens, and a few harbouring high worm burdens, generating mechanistically the classic overdispersed distribution that is a hallmark of helminth epidemiology. The dynamics of L1*,* L2*,* and L3 larvae within vectors are modelled deterministically, with differential equations defining their progression. The extrinsic incubation period of the parasite within blackflies takes approximately one week under tropical conditions. The dynamics of microfilariae within humans are also modelled deterministically. Density-dependent processes are assumed to operate upon parasite establishment within humans and blackfly vectors, vector survival (affected by the number of ingested microfilariae), and the probability that a female worm is mated.

**Supplementary Methods 1.** Modelling ivermectin and moxidectin treatment effects on *Onchocerca volvulus* (5)

##### Microfilaricidal effect

The microfilaricidal effect was modelled as an excess per capita death rate of microfilariae following treatment, $\mu'_M(\tau_{(i)})$, as done by Basáñez et al. (6),

$$\mu'_M(\tau_{(i)}) = (\tau_{(i)} + \nu)^{-\varpi} \quad \text{[Eqn. S1]}$$

where parameter $\tau_{(i)}$ is the time since individual $i$ received treatment. This equation models the large initial finite microfilaricidal effect at the point of treatment, defined by $\nu$, followed by the decline in this excess rate as $\tau_{(i)}$ increases, controlled by $\varpi$. Upon a subsequent treatment, $\tau_{(i)}$ is reset to zero, and the equation returns to the large initial finite microfilaricidal rate before beginning the decay process again.

##### Embryostatic effect

The embryostatic effect is characterised by temporary sterilisation of adult female worms modelled as a treatment-induced excess per capita rate of fertile female worms becoming non-fertile, $\lambda'_M(\tau_{(i)})$. The terms non-fertile and infertile worms refer to two different processes, the former describing the natural cycles of fertility and non-fertility characteristic of *Onchocerca volvulus*, which leads to re-fertilisation upon a subsequent mating event (4), and the latter describing permanent sterility. The equation for the temporary embryostatic effect is given by (6),

$$\lambda'_M(\tau_{(i)}) = \lambda^{max} e^{(-\varphi \tau_{(i)})} \quad \text{[Eqn. S2]}$$

where $\lambda^{max}$ is the maximum rate of drug-induced embryostasis with parameter $\varphi$ controlling the rate of decay of the drug-induced embryostasis as the time from treatment, $\tau_{(i)}$, increases.

The parameters used for ivermectin were those previously estimated by Basáñez et al. (6). Turner et al. (7) fitted Eqns. S1 and S2 to the moxidectin Phase II clinical trial data (8) and estimated the corresponding parameters for moxidectin.

**Permanent sterilising effect**

Repeated doses of ivermectin were assumed to be associated with a cumulative (and potentially permanent) sterilising effect on adult worms, estimated at around 30–35% per dose (9). In EPIONCHO-IBM this effect applies from the second dose onwards. The Phase II and III clinical trials used single-dose moxidectin (8, 10, 11), so there is no available information on any potential cumulative effects of moxidectin on macrofilariae. However, by analogy between moxidectin and ivermectin, we assumed that moxidectin has the same 35% cumulative effect on adult female worms, per dose (from the second dose onwards). By way of sensitivity analysis, we also run simulations assuming that moxidectin may have a permanent sterilising effect (proportion of fertile female worms removed from the parasite population) twice as large as that of ivermectin (i.e., 70%) for moxidectin max, in virtue of its longer half-life in humans (20-43 days compared to approximately 1 day for ivermectin) (12). Supplementary Table 1 summarises the parameter values for the effects of ivermectin (IVM), moxidectin (MOX) and moxidectin max (MOX MAX) on *O. volvulus*.

When simulating IVM MDA, the eligible population was aged 5 years and older; when simulating MOX and MOX MAX MDA, the eligible population was aged 4 years and older (13, 14).

**Supplementary Table 1.** Parameter definitions and values for anti-parasitic effects of ivermectin (IVM), moxidectin (MOX) and moxidectin max (MOX MAX) used in EPIONCHO-IBM

| **Parameter or Variable** | **Definition** | **Value, Units and References** |
|---|---|---|
| $\mu'_M(\tau_{(i)}) = (\tau_{(i)} + v)^{-\varpi}$ | Microfilaricidal effect: Drug-induced per capita rate of excess mortality of *Onchocerca volvulus* microfilariae at time $\tau_{(i)}$ since treatment | Controlled by $v$ and $\varpi$, $yr^{-1}$ |
| $v$ | Constant to allow for very large yet finite microfilaricidal effect upon treatment with drug | IVM (6): 0.0096<br>MOX (7) & MOX MAX: 0.04 |
| $\pi$ | Shape parameter for excess mortality of microfilariae following treatment with drug | IVM (6): 1.25<br>MOX (7) & MOX MAX: 1.82 |
| $\lambda'_M(\tau_{(i)}) = \lambda^{max} e^{(-\varphi\ \tau(i))}$ | Embryostatic effect: Drug-induced per capita rate of reversion from fertile to non-fertile adult female worms at time $\tau_{(i)}$, since treatment | Controlled by $\lambda^{max}$ and $\varphi$, $yr^{-1}$ |
| $\lambda^{max}$ | Maximum rate of drug-induced female worm (transient) sterilisation | IVM (6): 32.4 $yr^{-1}$<br>MOX (7) & MOX MAX: 462 $yr^{-1}$ |
| $\varphi$ | Rate of decay of drug-induced female worm (transient) sterilisation | IVM (6): 19.6 $yr^{-1}$<br>MOX (7) & MOX MAX: 4.83 $yr^{-1}$ |
| $\lambda'_p$ | Irreversible sterilising effect: Proportion of adult female worms made permanently infertile at each treatment round (i.e., per dose, from second dose) | IVM (9): 0.345<br>MOX: 0.345 (assumed to be equal to ivermectin's) (9)<br>MOX MAX: 0.70 |

**Supplementary Methods 2.** Calculation of the (mean) number of years to reach < 1% microfilarial prevalence

For each IU, 200 parameter sets were simulated using EPIONCHO-IBM, and the outputs were aggregated by calculating the mean number of years to reach < 1% microfilarial prevalence for each of the 12 alternative treatment strategies (ATS) combinations (Table 4 of Main Text). Runs achieving this target before the start of the ATS period (2026 through to 2040), were assigned a value of 0 years, while those failing to reach the target by the end of 2040 were assigned a value of 16 years. As an example, and for 5 hypothetical simulations for a given IU and ATS combination, the first reaching the target in 2024, the second doing so in 2027, the third in 2030, the fourth in 2033, and the fifth failing to do so, the assigned values would, respectively, be 0, 2, 5, 8, and 16, yielding a mean number of years to reach < 1% mf prevalence of 6.2 years.

**Supplementary Table 2.** Policy-Relevant Items for Reporting Models in Epidemiology of Neglected Tropical Diseases (PRIME-NTD) summary table

For the analyses presented, we adhered to the Five Principles of the Neglected Tropical Diseases (NTD) Modelling Consortium for good practice in policy-relevant NTD modelling (15), and briefly describe below the five tenets, how they were fulfilled, and where in the Main Text and/or Supplementary File they can be found.

| **Principle** | **What has been done to satisfy the principle?** | **Where in the manuscript is this described?** |
|---|---|---|
| Stakeholder engagement | Discussions with collaborators, other modellers and funders | Acknowledgements and funding statement |
| Complete model documentation | Brief description of EPIONCHO-IBM provided; schematic representation of the model illustrated; details of drug effects on the parasite explained; references to full model description cited; model code is publicly available | Main Text: Section 2.1 (EPIONCHO-IBM); Section 2.2 (Modelling interventions); Section 2.4 (Fitting EPIONCHO-IBM to multiple-time point data); Supplementary Figure 1 Supplementary Methods; Supplementary Table 1; Code availability section; References of Main Text and Suplementary File 1 |
| Complete description of data used | Description of HISTONCHO dataset (16) outlined and reference cited | Main Text: Section 2.3 (Implementation units (IUs) and intervention histories); Data availability section; References of Main Text and Suplementary File 1 |
| Communicating uncertainty | 95% uncertainty intervals (2.5th-97.5th quantiles) of 200 stochastic simulations shown; distribution of the (mean) number of years necessary to reach < 1% microfilarial prevalence illustrated using box-plots | Main text Figure 2 and Figure 3 |
| Testable model outcomes | Data on multiple moxidectin MDA not yet available | Discussion and Conclusion |
